# An idiosyncratic, zonated stroma encapsulates desmoplastic liver metastases and originates from injured liver

**DOI:** 10.1101/2022.08.24.22279162

**Authors:** Carlos Fernández Moro, Sara Harrizi, Yousra Hamidi, Natalie Geyer, Danyil Kuznyecov, Evelina Tidholm-Qvist, Media Salmonson Schaad, Andrea C. del Valle, Sara Söderqvist, Lorand Bozóky, Ernesto Sparrelid, Luc Dirix, Peter B Vermeulen, Béla Bozóky, Jennie Engstrand, Marco Gerling

## Abstract

Colorectal cancer liver metastases (CRLM) grow in two major patterns defined by the histomorphology of the invasion front, *replacement* or *desmoplastic*. The desmoplastic pattern, in which a stromal rim separates tumor cells and liver parenchyma is a strong positive prognostic factor, implying favorable biological features. However, the origin of the perimetastatic stroma is unknown and the underlying biological mechanisms are unclear. Here, we created spatial growth pattern maps of resected CRLM at cell-level resolution using digital pathology and quantified growth pattern heterogeneity at unprecedented resolution. We manually generated > 60’ 000 individual digital annotations on 543 metastases from 263 consecutive patients. We found that, in contrast to standard growth pattern assessments, high-resolution scoring revealed the prognostic outcome to be dependent on growth pattern proportions, such that survival improved with increasing fractions of desmoplastic encapsulation. The desmoplastic pattern was coupled to decreased tumor viability and to preoperative chemotherapy, hinting at a potentially causative connection of tumor viability and fibrotic encapsulation. Analyses of the cellular constituents of the rim revealed previously unrecognized liver parenchymal remnants. Spatial quantitation of liver remnants in the rim uncovered its gradual zonation from benign-like fibrosis at the liver side to tumor-associated stroma at the metastasis edge. Together, our data suggest that the perimetatstic “desmoplastic” capsule primarily results from a reparative hepatic process in conjunction with inefficient tumor cell colonization of liver plates, rather than from actively induced desmoplasia. We posit a model in which efficient replacement-type growth that precludes a mature hepatic injury reaction determines prognosis. Our results underscore tumor-cell replacement of hepatocytes as key for liver metastatic progression and suggest that the spatial heterogeneity of tumor invasion can be leveraged to understand fundamental mechanisms of metastatic growth.

## Introduction

Tumor cell invasion into healthy tissue is a hallmark of cancer^1^. Malignant cells can use different anatomic trajectories to colonize healthy organs, as revealed by histological examination. For example, in liver metastases of gastrointestinal and breast cancers, tumor cells frequently invade the hepatic plates by replacing hepatocytes while co-opting the stromal scaffolds and the microvascular architecture^2^. This process is largely tolerated by the hepatic immune system, as reflected by the absence of a significant immune cell infiltrate at the invasion front^3^. In contrast, melanoma metastases can exploit intra- and periluminal spaces of the host vasculature for invasion, which is rarely seen in metastases from other primary tumors^2,4^. These tumor-specific invasion patterns highlight the role of distinct capabilities for metastatic invasion, which can be observed histopathologically. However, one growth pattern is shared by liver metastases of several primary tumor types, and is uniformly associated with a favorable prognosis: Metastases surrounded by a rim of fibrotic stroma have a more favorable outcome in colorectal, gastric, and pancreatic adenocarcinoma, as well as in melanoma^2,5–9^. It is generally assumed that encapsulated metastases grow by continuous expansion requiring angiogenesis, although mechanistic evidence for this model is lacking, and observational studies to support it have largely focused on the microvasculature^2,10^. Desmoplastic metastases may respond better to anti-angiogenic therapy than replacement-type metastases^8^, underscoring a potential role of angiogenesis specifically in the desmoplastic pattern. However, how angiogenesis might causally be linked to the pattern of tumor invasion is an open question.

Clinically, the distinct patterns of metastatic invasion are best studied in colorectal cancer liver metastases (CRLM) because surgical resection is the standard-of-care, which results in a high number of histological specimens^11^. Based on pathological evaluation and in analogy to the general modes of tumor invasion, the two major CRLM growth patterns have been systematized into *desmoplastic* and *replacement* patterns (**Figure 1a**)^2^. Desmoplastic-type metastases are surrounded by a rim of fibroblast-like stromal cells, extracellular matrix (ECM) deposits, and variable immune cell infiltrates, separating tumor cells from the liver parenchyma^2,8,12^. The replacement pattern is defined by direct physical contact between hepatocytes and invading tumor cells; it is assumed that in this pattern, invading tumor cells replace resident hepatocytes and co-opt the liver stromal architecture as well as its sinusoidal vasculature^2,8,13^. A third, rare pattern in which hepatocytes appear flattened around an expanding tumor mass without distinct tumor-hepatocyte contact is termed *pushing*^2^. The more favorable outcome of desmoplastic CRLM has been demonstrated in numerous retrospective studies^5,14,15^.

**Figure 1.**
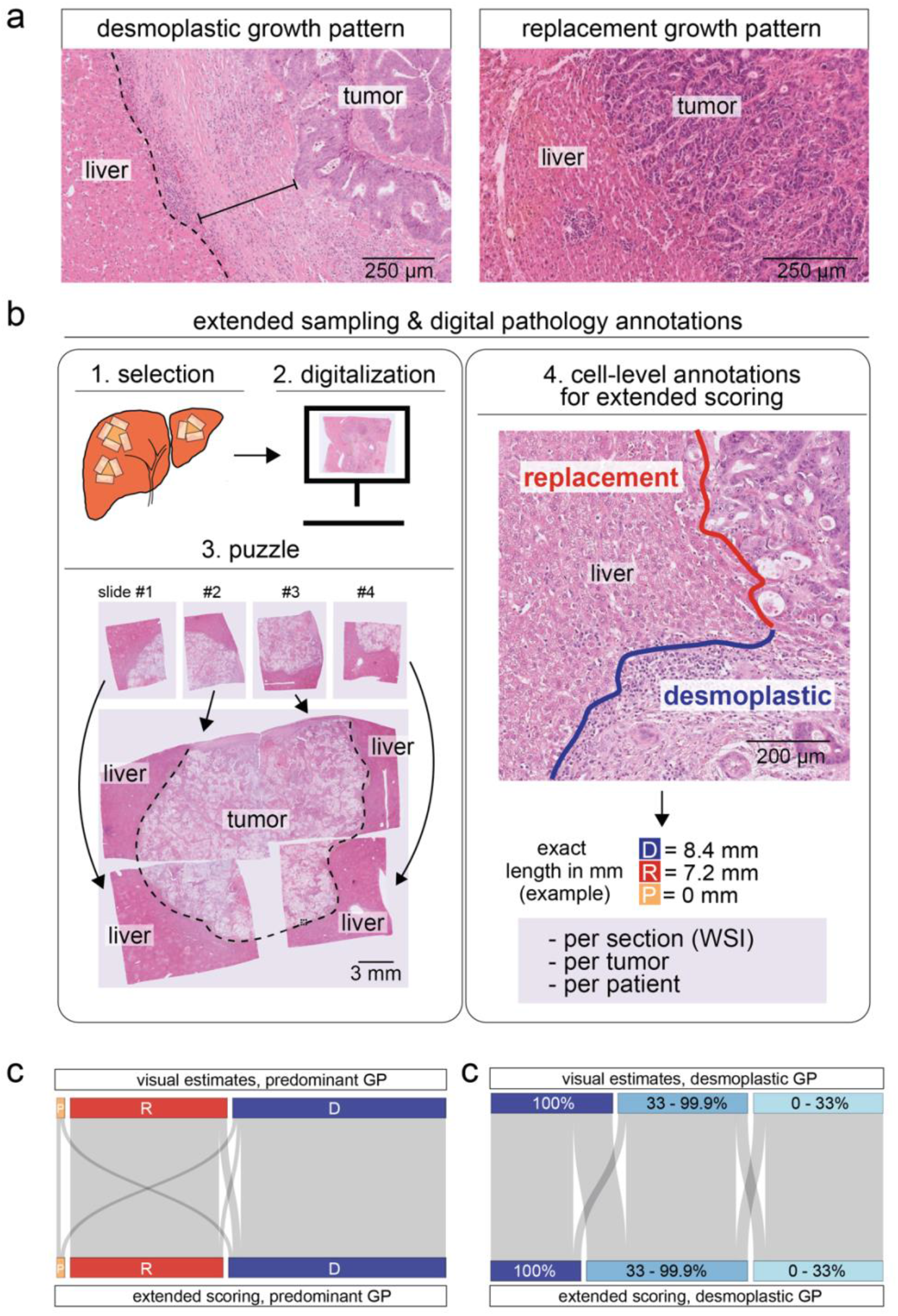
Extended *versus* standard scoring of colorectal cancer liver metastases. **a)** *Desmoplastic* metastases (left panel): a stromal rim separates tumor cells and liver parenchyma. *Replacement* metastases: cancer are in contact with hepatocytes, which they appear to replace. **b)** Schematic of extended annotations on whole slide images (WSIs). 1 (<u>selection</u>): All sections representing the largest metastasis diameter are selected. 2 (<u>digitalization</u>): Those slides of the largest diameter maximizing representation of the circumference along the tumor-liver interface are digitized, overlapping slides are disregarded (not shown). 3 (<u>puzzle</u>): Several WSI from selected slides are combined. 4: (<u>annotation</u>): Each slide is annotated manually for each growth pattern, indicated in different colors. Dashed black line indicates tumor-liver-interface. **c)** Sankey plot of reclassifications resulting from visual estimates vs. extended WSI-based scoring. **d)** Sankey plot of reclassifications from the indicated percentages of desmoplastic encapsulation; n = 97 patients for (c) and (d)

Despite their prognostic value, the molecular bases for the emergence of the different patterns remain unknown. The etiology of the desmoplastic pattern is particularly enigmatic, as it is unclear whether tumors actively induce the formation of the perimetastatic stroma, or if the capsule is the result of a liver-specific reaction, uncoupled from tumoral factors^2^. No correlations between the growth patterns and oncogenic *KRAS, BRAF*, and *NRAS* mutations, which characterize aggressive colorectal cancer subtypes, have been found^15,16^. Microsatellite instability (MSI) is more frequent in desmoplastic than in replacement-type metastases; however, the absolute frequencies are reportedly low in both (14.6% in desmoplastic vs. 3.6% in replacement-type metastases, according to a recent study)^15^ and hence, MSI does not sufficiently explain growth pattern biology.

Desmoplasia of the tumor stroma is commonly regarded as the result of resident fibroblast activation, driven by tumoral factors and reinforced by the recruitment of other cellular sources; in the prevailing model, desmoplasia is seen either as an active damage-response of the preexisting stroma to the presence of tumor cells, or as a result of tumor cell-derived factors actively shaping their stromal niche^17^. Although certain marker proteins, such as alpha-smooth muscle actin (ASMA) and different collagens in the ECM characterize cancer-associated desmoplasia, regardless of its cellular and mechanistic origin^18–20^, none of these markers is specific^17,19^.

Here, we sought a deeper understanding of the origin and development of the desmoplastic rim in CRLM. We provide extensive digital annotations, reflecting the heterogeneity of the capsule formation at the intra- and interlesional levels, connected to clinical outcomes. In contrast to results from standard visual scoring^2^, we find a strong direct relationship between outcome and growth pattern proportion, such that the risk of local relapse and death decreased with an increase in the proportion of fibrotic encapsulation. Quantitative spatial analyses of the fibrotic rim suggested that perimetastatic fibrosis evolves from the liver parenchyma towards the metastasis. Importantly, we discover and quantify remnants of portal tracts in the rim and the tumor center of desmoplastic-type metastases, providing novel, direct evidence that the stromal rim largely represents atrophic liver.

Together, our data support a model in which the desmoplastic rim around liver metastases results from a failed replacement-type tumor invasion into the liver parenchyma with subsequent liver fibrosis analogous to inflammatory and atrophic liver disease.

## Results

### Extended sampling and growth pattern annotations reveal fine-grained prognostic strata

Conventionally, growth pattern fractions are estimated by visual assessment of the tumor-liver interface on standard hematoxylin & eosin (H&E) stained sections, as defined in the international consensus guidelines^2^. Using this scoring approach, only those CRLM that are fully encapsulated by fibrotic stroma have a significantly better prognosis than metastases with even the smallest fraction of replacement growth^15,21^. Consequently, the current clinical guidelines recommend classifying only fully encapsulated metastases as desmoplastic^2^. This cut-off of 100% desmoplastic growth was derived from testing different fractions of encapsulation in large patient cohorts from multiple clinical centers, in which only fully encapsulated metastases showed favorable outcomes^2,15,21^. The resulting scoring algorithm is clinically applicable, highly reproducible, and identifies patients with excellent prognoses. However, mechanistically, it suggests that replacement growth determines outcome regardless of its proportion over the whole length of the invasion front. This posits a deterministic Boolean model, which is difficult to reconcile with the probabilistic character of biological systems; a probabilistic model would predict survival to increase in a, e.g. linear or logarithmic manner with respect to the extent of a positive physical trait, in this case, the proportion of the desmoplastic pattern.

Therefore, we sought to test different cut-offs for the proportion of desmoplastic encapsulation for their impact on prognostication after addressing the two main shortcomings of the current scoring methodology: (1) the extent of sampling (i.e., an extensive representation of the invasion front), and (2) the resolution of growth pattern scoring (**Figure 1b**). First, we systematically and extensively sampled the invasion front of all metastases in each patient, approximating a complete panoramic central slice along the largest tumor diameter (**Figure 1b**)^22^. This was possible due to highly standardized sampling introduced in 2012 at the Department of Clinical Pathology and Cancer Diagnostics, Karolinska University Hospital (KUH), Stockholm, Sweden. Secondly, we used digital whole slide images (WSIs) of all resulting sections to manually annotate the growth patterns over the entire invasion front for each slide (**Figure 1b**). Our cohort comprised all consecutive patients undergoing their first operation for CRLM between May 2012 and December 2015 at Karolinska University Hospital, Stockholm, Sweden (*n* = 263 patients, **Table 1**).

**Table 1:**
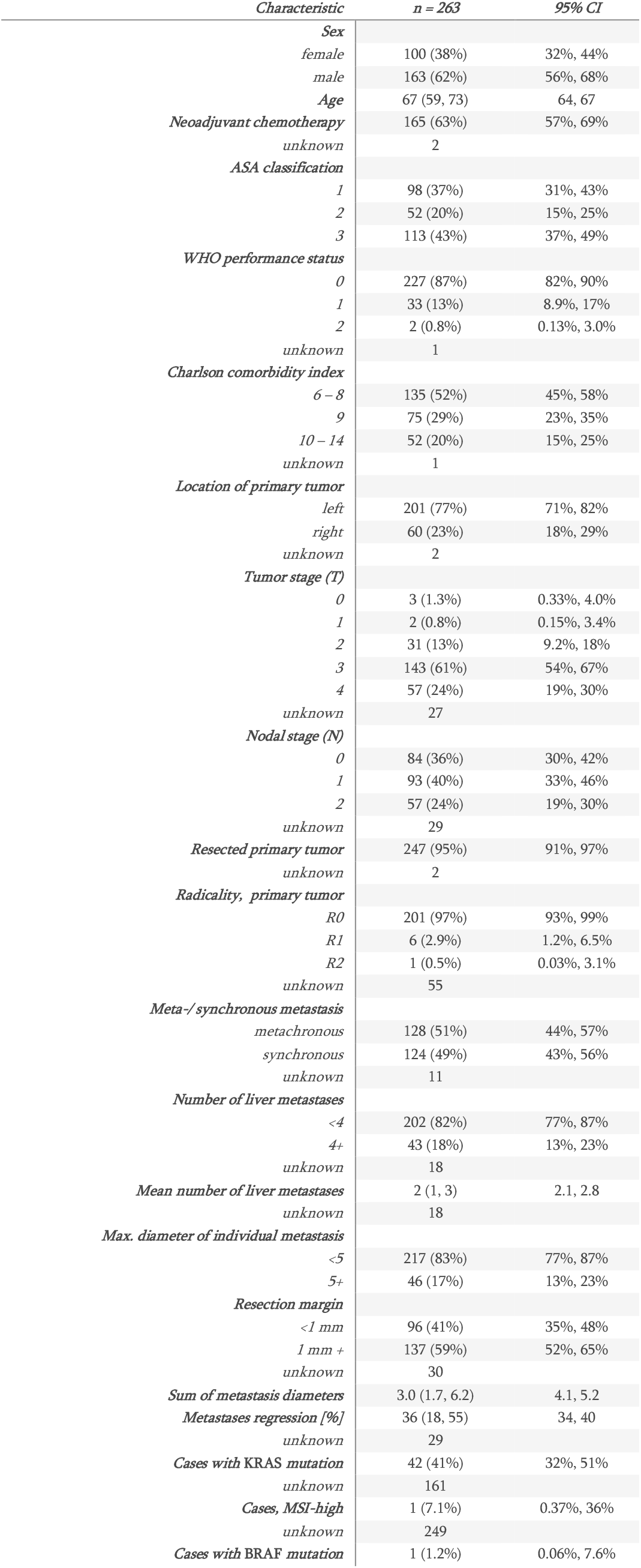
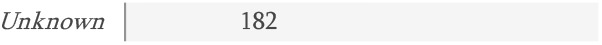
Clinical characteristics of all patients in the cohort. Column 1: number of patients or median with interquartile rangein brackets. Column 2, CI: confidence interval. ASA: American Society of Anesthesiologists, WHO: world health organization, KRAS: Kirsten rat sarcoma virus gene, MSI: microsatellite instability, BRAF: B-Raf Proto-Oncogene

| *Characteristic* | *n = 263* | *95% CI* |
| --- | --- | --- |
| **Sex** |  |  |
| female | 100 (38%) | 32%, 44% |
| male | 163 (62%) | 56%, 68% |
| **Age** | 67 (59, 73) | 64, 67 |
| **Neoadjuvant chemotherapy** | 165 (63%) | 57%, 69% |
| unknown | 2 |  |
| **ASA classification** |  |  |
| 1 | 98 (37%) | 31%, 43% |
| 2 | 52 (20%) | 15%, 25% |
| 3 | 113 (43%) | 37%, 49% |
| **WHO performance status** |  |  |
| 0 | 227 (87%) | 82%, 90% |
| 1 | 33 (13%) | 8.9%, 17% |
| 2 | 2 (0.8%) | 0.13%, 3.0% |
| unknown | 1 |  |
| **Charlson comorbidity index** |  |  |
| 6 – 8 | 135 (52%) | 45%, 58% |
| 9 | 75 (29%) | 23%, 35% |
| 10 – 14 | 52 (20%) | 15%, 25% |
| unknown | 1 |  |
| **Location of primary tumor** |  |  |
| left | 201 (77%) | 71%, 82% |
| right | 60 (23%) | 18%, 29% |
| unknown | 2 |  |
| **Tumor stage (T)** |  |  |
| 0 | 3 (1.3%) | 0.33%, 4.0% |
| 1 | 2 (0.8%) | 0.15%, 3.4% |
| 2 | 31 (13%) | 9.2%, 18% |
| 3 | 143 (61%) | 54%, 67% |
| 4 | 57 (24%) | 19%, 30% |
| unknown | 27 |  |
| **Nodal stage (N)** |  |  |
| 0 | 84 (36%) | 30%, 42% |
| 1 | 93 (40%) | 33%, 46% |
| 2 | 57 (24%) | 19%, 30% |
| unknown | 29 |  |
| **Resected primary tumor** | 247 (95%) | 91%, 97% |
| unknown | 2 |  |
| **Radicality, primary tumor** |  |  |
| R0 | 201 (97%) | 93%, 99% |
| R1 | 6 (2.9%) | 1.2%, 6.5% |
| R2 | 1 (0.5%) | 0.03%, 3.1% |
| unknown | 55 |  |
| **Meta-/synchronous metastasis** |  |  |
| metachronous | 128 (51%) | 44%, 57% |
| synchronous | 124 (49%) | 43%, 56% |
| unknown | 11 |  |
| **Number of liver metastases** |  |  |
| <4 | 202 (82%) | 77%, 87% |
| 4+ | 43 (18%) | 13%, 23% |
| unknown | 18 |  |
| **Mean number of liver metastases** | 2 (1, 3) | 2.1, 2.8 |
| unknown | 18 |  |
| **Max. diameter of individual metastasis** |  |  |
| <5 | 217 (83%) | 77%, 87% |
| 5+ | 46 (17%) | 13%, 23% |
| **Resection margin** |  |  |
| <1 mm | 96 (41%) | 35%, 48% |
| 1 mm + | 137 (59%) | 52%, 65% |
| unknown | 30 |  |
| **Sum of metastasis diameters** | 3.0 (1.7, 6.2) | 4.1, 5.2 |
| **Metastases regression [%]** | 36 (18, 55) | 34, 40 |
| unknown | 29 |  |
| **Cases with KRAS mutation** | 42 (41%) | 32%, 51% |
| unknown | 161 |  |
| **Cases, MSI-high** | 1 (7.1%) | 0.37%, 36% |
| unknown | 249 |  |
| **Cases with BRAF mutation** | 1 (1.2%) | 0.06%, 7.6% |

This cohort reflects most modern CRLM therapies, including monoclonal antibodies targeting vascular endothelial growth factor (VEGF) and epidermal growth factor receptor (EGFR), and at the same time allows for a long follow-up time.

For the first *n* = 97 patients, we assessed the growth patterns both visually and with our WSI-based approach to evaluate the impact of extended, cell-level scoring on growth pattern classifications. As expected, scores from digital annotations were more granular, while standard visual scoring was often performed in 5% – 10% increments (**Supplementary Figure 1a**). Overall, we found good agreement between digital annotations and visual assessments performed by the same raters with more than one year gap between the assessments (ET, DK, Cohen’s Kappa [predominant pattern] = 0.82), or when compared to external expert visual assessment (PV, who coined the growth patterns more than two decades ago^13^ ; Cohen’s Kappa [predominant pattern] = 0.72). Visual estimates deviated from annotation-based scoring particularly at the extremes, towards 0% and 100% (**Supplementary Figure 1b**). Next, we assessed whether visual estimates and digital annotations changed the classification of each patient into the categories of their predominant growth patterns. For most patients, the predominant growth pattern classification remained unchanged, while n = 5 patients (5 %) were reclassified (**Figure 1c**). However, using different cut-offs for desmoplastic encapsulation as previously suggested (<33%, 33–<100%, and 100% desmoplastic pattern)^21^, n = 18 patients (19%) were reclassified (**Figure 1d**). When we used the predominant pattern to define the strata, which previously demonstrated a strong association with prognosis in numerous studies^5,14^, we found that both scoring methods identified desmoplastic metastases to be associated with a more favorable overall survival (OS), as expected (**Supplementary Figure 1c**). Applying different cut-offs for desmoplastic encapsulation, we strikingly observed a trifurcation of the strata with a progressively improving outcome from <33% over 33–<100% to 100% desmoplastic encapsulation (**Supplementary Figures 1c and d**); the trifurcation was most distinct in patients that had received neoadjuvant chemotherapy, and when using extended scoring (**Supplementary Figures e and f**). These data suggested that enforced detailed assessment of the growth patterns by the same raters affects the granularity of growth pattern assessments, which alters the classification of some cases and which might improve prognostic stratification.

The slight, but potentially important differences between standard and extended scoring prompted us to completed digital annotations for the entire cohort comprising *n* = 263 CRLM patients, for which a total of *n* = 897 WSIs were evaluated, resulting in 60’ 878 individual annotations. For *n* = 231 patients, a predominant growth pattern could be determined, after excluding patients with complete regression whose growth patterns were not estimated according to the guidelines^2^, and those patients with no available representative sections. In the final series, n = 122 (53%) patients had metastases with predominantly desmoplastic growth ^5^, n = 105 (45%) had predominantly replacement growth, and n = 4 (2%) patients had predominantly pushing metastases, which is in line with previously reported distributions^2,14^. Over the entire cumulative circumference of all tumors, the desmoplastic pattern was the major pattern (55%), followed by replacement (43%) and pushing (2%). As expected, CRLM with a predominantly desmoplastic growth pattern had a significantly better OS and liver-specific progression-free survival (hepatic-PFS, hPFS) compared to predominantly replacement-type metastases, and to rare cases of predominantly pushing metastases (**Figure 2a**).

**Figure 2:**
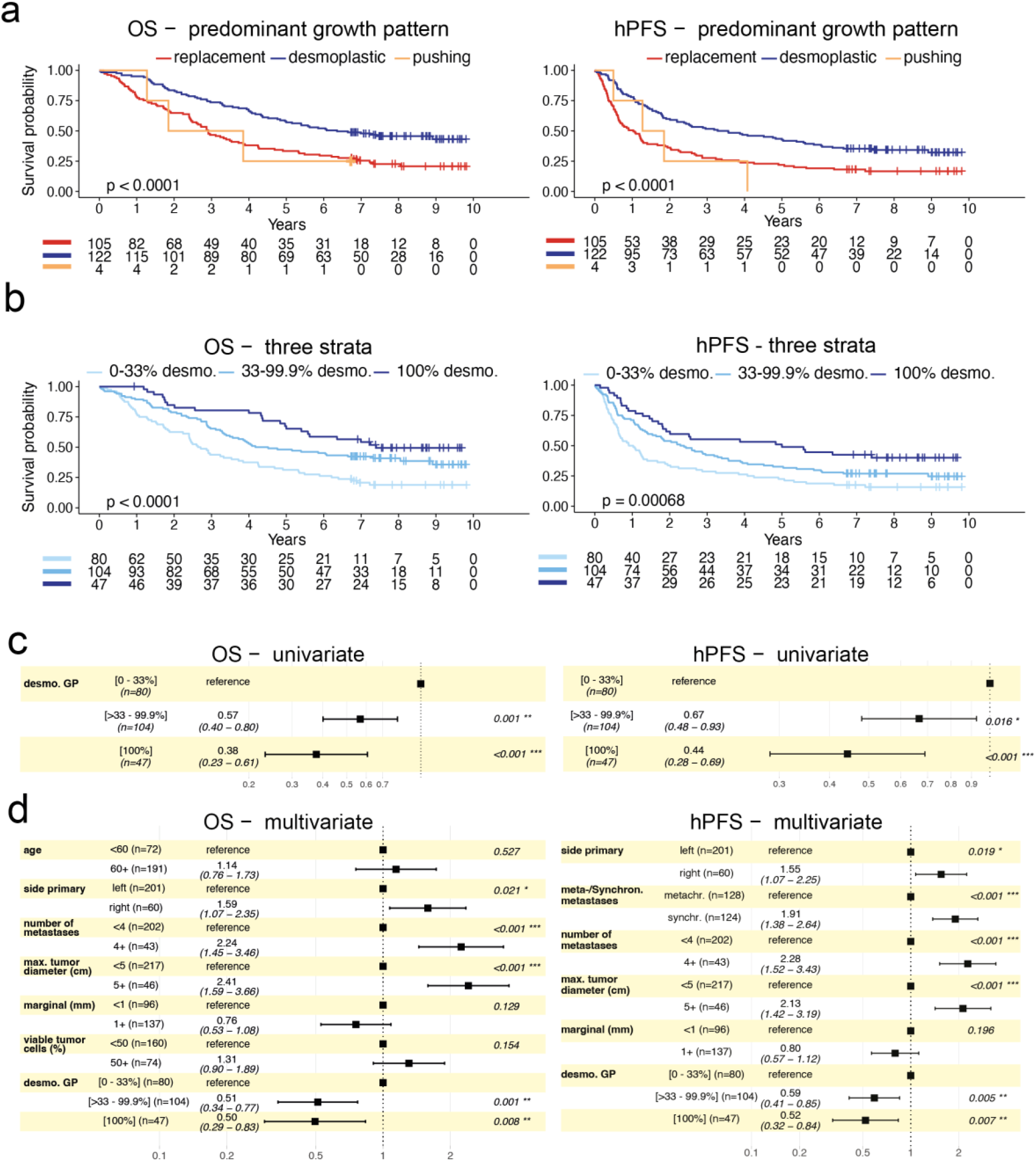
Prognostic trifurcation dependends on the fraction of desmoplastic encapsulation. **a)** Overall survival (OS, left) and liver-specific (hepatic) progression-free survival (hPFS, right), when scored by the predominant growth pattern: replacement, desmoplastic, and pushing. **b)** OS and hPFS stratified by the indicated proportions of desmoplastic encapsulation. Log-rank p-values given in the plots, (a) and (b). **c)** Univariate cox proportional hazard regression using three strata of desmoplastic encapsulation, OS left; hPFS right. **d)** Multivariate cox proportional hazard analysis according using desmoplastic encapsulation in three strata; OS left; hPFS right. Hazard ratio (HR) with 95% confidence intervals in brackets. Wald-test p-value, asterisks indicate significance levels, ***: p < 0.001, **: p < 0.01, *: p < 0.05.

The extensive digital annotations allowed further stratification of growth pattern fractions. Leveraging high-resolution digital scoring revealed a strong growth pattern proportion-dependency of outcome, such that decreasing proportions of desmoplastic encapsulation were associated with increasingly worse OS and hPFS (**Figure 2b**). This finding was in contrast to previous results based solely on visual estimates that suggested a Boolean behavior, where any evidence of replacement growth placed patients in the group with an equally bad outcome compared to fully desmoplastic metastases^2,21^. Uni- and multivariate analyses of three strata for desmoplastic encapsulation (low: 0–33%, medium: 33–<100%, high: 100%) in our cohort confirmed longer OS and hPFS in both medium (hazard ratios [HR] = 0.57 and 0.67 respectively) and high (HR = 0.38 and 0.44 respectively) proportion strata (**Figures 2c and d**).

Hence, the degree of desmoplastic encapsulation, rather than the mere presence of non-desmoplastic growth, has prognostic value for CRLM patients after surgery.

### Increased desmoplastic encapsulation after neoadjuvant chemotherapy

A recent study found an increased frequency of desmoplastic metastases in patients who received neoadjuvant chemotherapy^16^. However, previous studies have not reported similar findings (systematic review, ref.^14^). From a tumor biology perspective, a growth pattern switch after chemotherapy could mechanistically connect decreased tumor cell fitness, resulting from cytotoxic therapy, with perimetastatic fibrosis. Hence, we asked whether we could detect differences in growth pattern frequency after chemotherapy using our extended scoring. We also found that the frequency of desmoplastic metastases was significantly higher in patients who had received neoadjuvant chemotherapy than in those who had not (**Figure 3a**). Our annotation data further allowed us to quantify the degree of encapsulation on each slide of each metastasis for every patient, which confirmed the increase in desmoplastic encapsulation after chemotherapy (**Figure 3b**).

**Figure 3:**
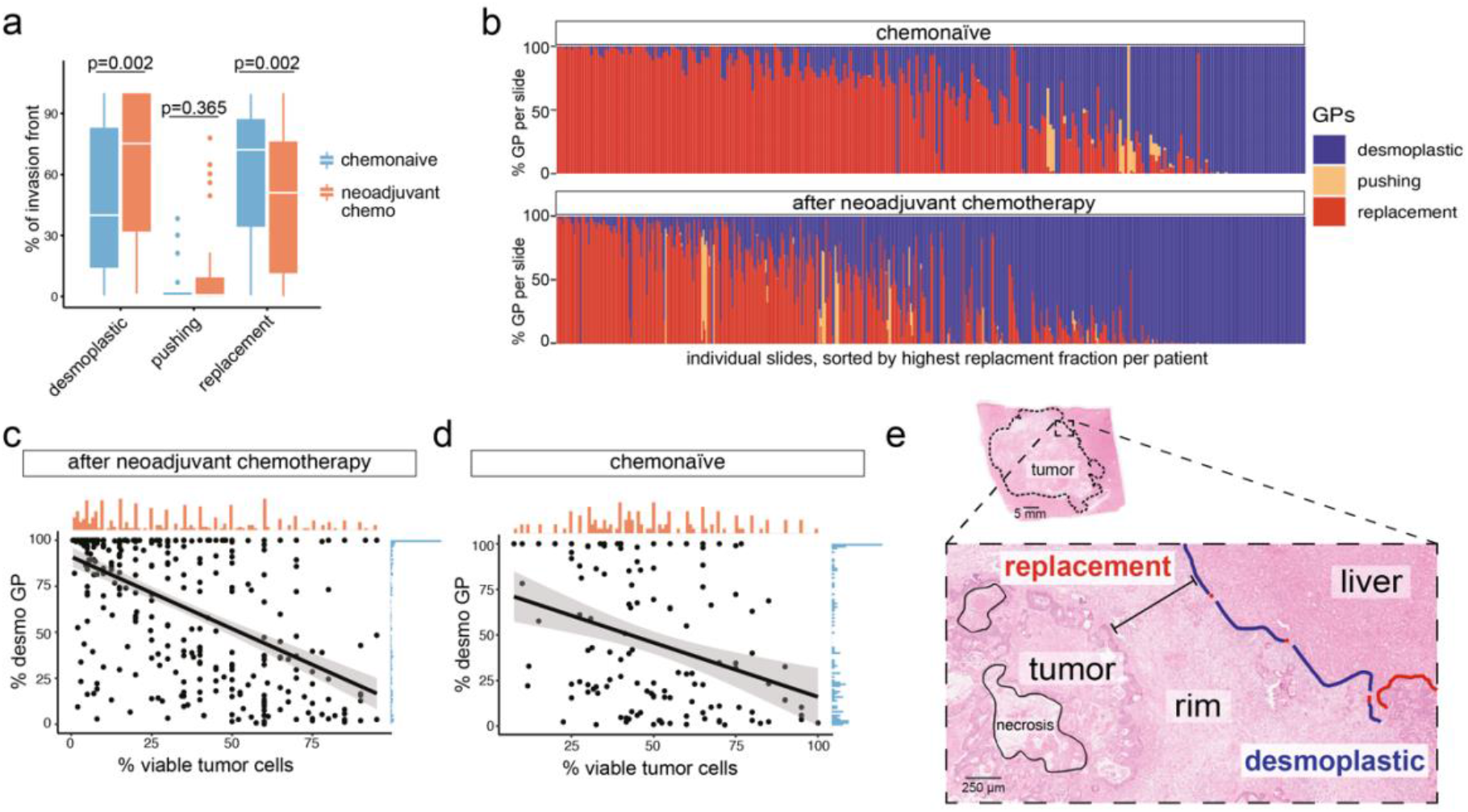
Impact of neoadjuvant chemotherapy on growth pattern frequency. **a)** Change in percentages of desmoplastic, pushing, and replacement annotations in patients who received chemotherapy vs. those who did not. **b)** Distribution of growth patterns per slide in chemo naïve patients and patients who received neoadjuvant therapy. Note the increase of desmoplastic encapsulation (blue). **c)** and **d)** Correlation of annotated percentage desmoplastic growth pattern and percentage of viable tumor cells in chemotherapy-treated **(c)** vs. untreated **(d)** patients. **e)** Representative example of a treated metastasis, illustrating viable tumor cells remote from the surrounding liver parenchyma. Black circled structures denote necrotic tumor, blue line the desmoplastic growth pattern, red line the replacement growth pattern.

Within the patient group that had received neoadjuvant treatment, there was a strong negative correlation between the fraction of viable tumor cells and the degree of desmoplastic encapsulation (Spearman’s r = -0.52, p = 3.5e-11, **Figure 3c**), while this association was weaker in the chemo naïve group (Spearman’s r = -0.24, p = 0.028, **Figure 3d**). Inspection confirmed that the remaining viable tumor cells were located both in regions of fibrotic encapsulation and in those with replacement-type growth (**Figure 3e**).

These results corroborate recent data suggesting that the fibrotic capsule can be induced by chemotherapy^16^. However, since the growth patterns cannot be determined preoperatively^2^, there is a risk of selection bias in our cohort. Except for a few individual cases, including young patients with limited disease, at our center surgical resection is not performed when progression is noted radiologically during neoadjuvant chemotherapy. Hence, metastases that progress upon chemotherapy are largely missing from the neoadjuvant group, which could skew growth pattern frequency towards the desmoplastic type. On the other hand, chemotherapy is sometimes omitted in patients with small singular metastases. When we compared clinicopathological characteristics of patients receiving neoadjuvant treatment to those who did not, we found that patients who had received chemotherapy were younger and had significantly more metastases than chemo naïve patients. However, the combined sum of tumor diameters was similar, indicating comparable tumor burden in both groups (**Table 2**).

**Table 2:** Comparison of patients that received chemotherapy and those that did not. Columns 1 and 2: Number of patients or median with interquartile range in brackets. Column 3: Results from Pearson’s Chi-squared test, Wilcoxon rank sum test, and Fisher’s exact test, where applicable. ASA: American Society of Anesthesiologists, WHO: world health organization, KRAS: Kirsten rat sarcoma virus gene, MSI: microsatellite instability, BRAF: B-Raf Proto-Oncogene

| Variable | n | Neoadjuvant chemotherapy |  | p-value<sup>2</sup> |
| --- | --- | --- | --- | --- |
|  |  | No, N = 96<sup>1</sup> | Yes, N = 165<sup>1</sup> |  |
| **Sex** | 261 |  |  | 0.004 |
| female |  | 26 (27%) | 74 (45%) |  |
| male |  | 70 (73%) | 91 (55%) |  |
| **Age** | 261 | 72 (63, 78) | 66 (57, 71) | <0.001 |
| **ASA classification** | 261 |  |  | <0.001 |
| 1 |  | 22 (23%) | 76 (46%) |  |
| 2 |  | 15 (16%) | 37 (22%) |  |
| 3 |  | 59 (61%) | 52 (32%) |  |
| **WHO performance status** | 260 |  |  | <0.001 |
| 0 |  | 69 (72%) | 156 (95%) |  |
| 1 |  | 25 (26%) | 8 (4.9%) |  |
| 2 |  | 2 (2.1%) | 0 (0%) |  |
| unknown |  | 0 | 1 |  |
| **Charlson comorbidity index** | 260 |  |  | <0.001 |
| 6 - 8 |  | 32 (33%) | 103 (63%) |  |
| 9 |  | 26 (27%) | 48 (29%) |  |
| 10 - 14 |  | 38 (40%) | 13 (7.9%) |  |
| unknown |  | 0 | 1 |  |
| **Location of primary tumor** | 259 |  |  | 0.8 |
| left |  | 74 (78%) | 125 (76%) |  |
| right |  | 21 (22%) | 39 (24%) |  |
| unknown |  | 1 | 1 |  |
| **Tumor stage (T)** | 235 |  |  | 0.6 |
| 0 |  | 0 (0%) | 3 (2.0%) |  |
| 1 |  | 0 (0%) | 2 (1.4%) |  |
| 2 |  | 11 (13%) | 20 (14%) |  |
| 3 |  | 57 (66%) | 85 (57%) |  |
| 4 |  | 19 (22%) | 38 (26%) |  |
| unknown |  | 9 | 17 |  |
| **Nodal stage (N)** | 233 |  |  | 0.8 |
| 0 |  | 31 (36%) | 52 (35%) |  |
| 1 |  | 36 (42%) | 57 (39%) |  |
| 2 |  | 19 (22%) | 38 (26%) |  |
| unknown |  | 10 | 18 |  |
| **Resected primary tumor** | 259 | 93 (97%) | 152 (93%) | 0.2 |
| unknown |  | 0 | 2 |  |
| **Radicality, primary tumor** | 207 |  |  | 0.6 |
| R0 |  | 77 (96%) | 123 (97%) |  |
| R1 |  | 2 (2.5%) | 4 (3.1%) |  |
| R2 |  | 1 (1.3%) | 0 (0%) |  |
| unknown |  | 16 | 38 |  |
| **Meta-/ synchronous metastasis** | 250 |  |  | <0.001 |
| metachronous |  | 66 (71%) | 61 (39%) |  |
| synchronous |  | 27 (29%) | 96 (61%) |  |
| unknown |  | 3 | 8 |  |
| **Number of liver metastases** | 243 |  |  | <0.001 |
| <4 |  | 85 (94%) | 115 (75%) |  |
| 4+ |  | 5 (5.6%) | 38 (25%) |  |
| Unknown |  | 6 | 12 |  |
| **Mean number of liver metastases** | 243 | 1 (1, 2) | 2 (1, 3) | <0.001 |
| Unknown |  | 6 | 12 |  |
| **Max. diameter of individual metastasis** | 261 |  |  | 0.064 |
| <5 cm |  | 74 (77%) | 142 (86%) |  |
| 5 cm |  | 22 (23%) | 23 (14%) |  |
| **Resection margin** | 231 |  |  | 0.5 |
| <1 mm |  | 38 (44%) | 58 (40%) |  |
| 1 mm + |  | 48 (56%) | 87 (60%) |  |
| unknown |  | 10 | 20 |  |
| **Sum of all metastasis diameters (cm)** | 261 | 3.0 (2.0, 6.5) | 3.0 (1.4, 6.0) | 0.3 |
| **Metastases regression [%]** | 232 | 46 (35, 60) | 28 (9, 46) | <0.001 |
| unknown |  | 12 | 17 |  |
| **Cases with KRAS mutation** | 102 | 4 (25%) | 38 (44%) | 0.2 |
| unknown |  | 80 | 79 |  |
| **Cases, MSI-high** | 14 | 0 (0%) | 1 (11%) | >0.9 |
| unknown |  | 91 | 156 |  |
| **Cases with BRAF mutation** | 81 | 1 (7.1%) | 0 (0%) | 0.2 |
| unknown |  | 82 | 98 |  |

### Presence of liver parenchymal remnants in the desmoplastic rim

If chemotherapy can convert replacement-type growth into perimetastatic desmoplasia, the resulting stromal rim is unlikely to be actively induced by tumor-intrinsic factors. Instead, perimetastatic fibrosis could represent a reaction of the liver parenchyma to cellular damage, for example, as a consequence of cytotoxic treatment. This model for the emergence of the capsule would predict the presence of liver parenchymal remnants in the rim. However, we only rarely observed mature hepatocytes in the rim by conventional light microscopy. Therefore, we used mRNA in situ hybridization (ISH) for the hepatocyte marker, albumin (*ALB*), to assess the presence of hepatocyte-related gene expression in the rim. Although ISH did reveal *ALB*-expressing cells in the perimetastatic capsule, these cells were smaller than normal hepatocytes and had lost their typical cuboidal morphology, likely reflecting regenerative changes (**Figure 4a**).

**Figure 4:**
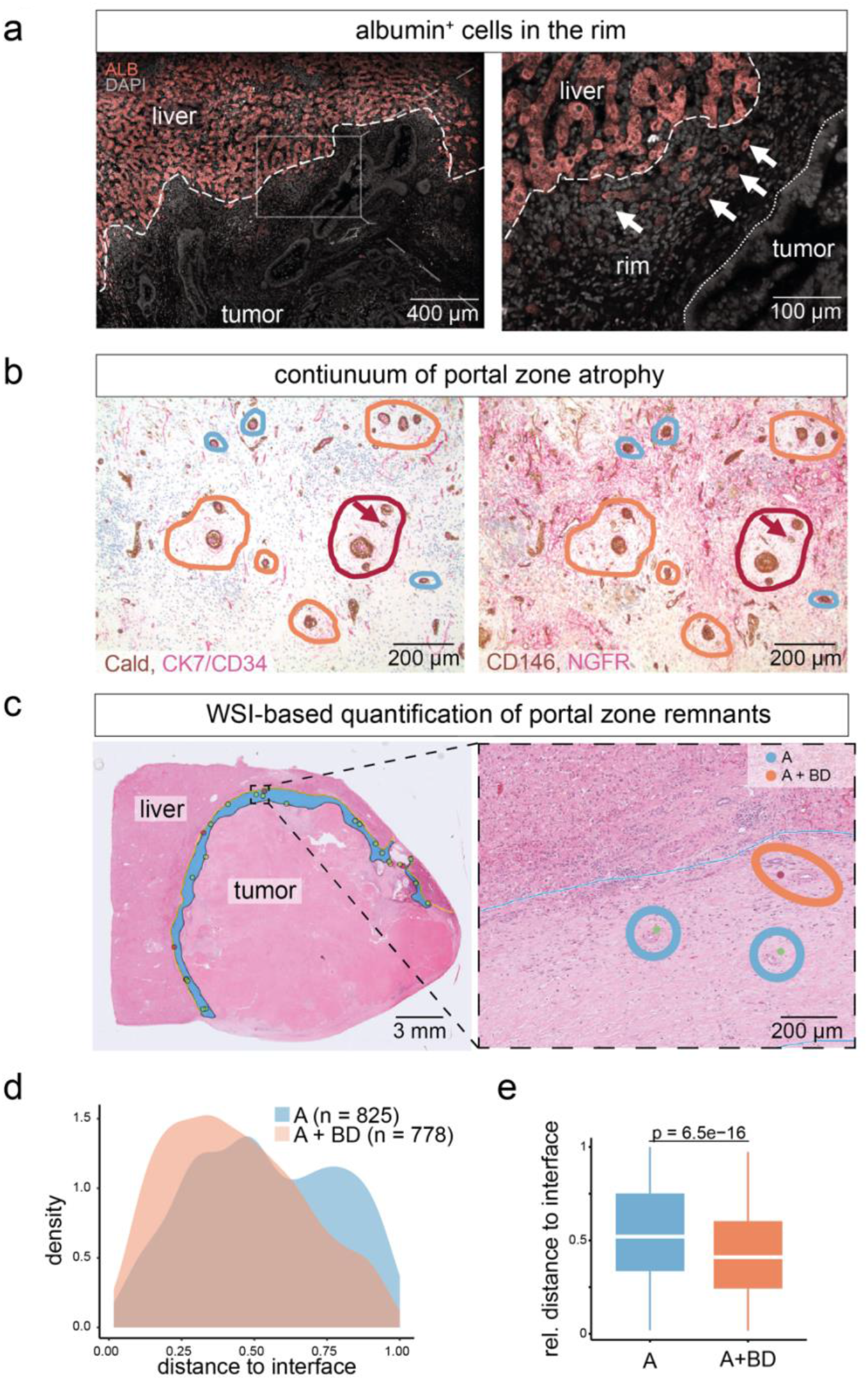
Liver parenchymal remnants in the desmoplastic rim. **a)** mRNA in situ hybridization of a desmoplastic CRLM for albumin (*ALB*), nuclear stain with DAPI (4′,6-diamidino-2-phenylindole). Arrows: *ALB+* cells in the rim. Dashed line: tumor-rim interface; dotted line: Tumor cell border towards the liver. Representative image of n = 4 desmoplastic metastases. **b)** Different degrees of portal tract (PT) atrophy in CLRM indicated by different colors: Red: scant porta zones with preserved branches of the hepatic artery, bile duct (arrows), and variably portal stroma. Orange: remnants of porta zones with preserved branches of the hepatic artery and variably portal stroma. Blue: most atrophic remnants, where only branches of the hepatic artery remain, without distinctive surrounding portal stroma. **c)** Whole-slide image (WSI) annotated for the desmoplastic rim and portal tracts: yellow line marks border of the desmoplastic rim and the liver; blue area marks the rim; red dots mark PTs consisting of artery and bile duct (A+BD), green dots mark PTs consisting of artery remnants (A). **d)** PT density in the desmoplastic rim per desmoplastic metastases (>95% desmoplastic, n=49 patients. **e)** Box plot of the distributions of PT arteries and PT remnants consisting of arteries and bile ducts, results from Wilcoxon rank sum test are presented in the panel. Cald: Caldesmon. CK7: cytokeratin 7, CD34: cluster of differentiation 34.

Liver lobules, the repetitive functional units that make up most of the liver parenchyma, contain portal triads (PTs) at their corners. PTs include branches of three structures, (*1*) the portal vein, (*2*) the hepatic artery, and (*3*) the bile duct. PTs are hubs for fibrosis development in non-malignant fibrotic conditions, including periportal fibrotic expansion in chronic hepatitis and primary biliary diseases, as well as parenchymal scarring following hepatocyte atrophy^23^ and necrosis^24^. PTs or their remnants display different degrees of atrophy, embedded in the fibrotic stroma in areas where hepatocytes are no longer detectable^25^. The presence of PTs in the center of replacement-type metastases has been described previously and supports the generally accepted model where tumor cells grow between portal structures by replacing the bulk of hepatocytes^2^. During clinical routine, we observed different degrees of PT atrophy. Aside from full triads, PT remnants with the remaining artery and bile duct (A+BD) can be seen, which are devoid of the (then obliterated) portal vein, as well as those with only the artery (A) remaining. Such single arteries are often found embedded in a sheet of distinct, portal stroma with variably preserved expression of the portal stroma markers, NGFR and CD34 (**Figure 4b**)^2^, but can also be identified based on H&E stains by the characteristic morphology of the surrounding stroma.

Therefore, to test the hypothesis that the perimetastatic fibrotic stroma of desmoplastic-type metastases is a remnant of injured liver plates, we revisited those cases in our cohort with >95% desmoplastic encapsulation (*n* = 49 cases, corresponding to *n* = 141 WSIs) and used H&E stained WSIs to identify remnants of PTs in the desmoplastic rim (**Figure 4c**). Strikingly, quantitation revealed PT remnants in the fibrotic rim in all cases, although at varying frequency (**Supplementary Figure 2a**). We also found similar degrees of PT atrophy in primary hepatic tumor types, suggesting that the continuum of atrophy is not specific to metastases (**Supplementary Figures 2b**).

Given the sequence of the loss of anatomical structures (from full triads over A+BD remnants to A remnants; **Figure 4b and Supplementary Figure 2b**), A+BD type PTs arguably represent an earlier stage of PT atrophy than A remnants. When we analyzed the spatial distribution of A and A+BD PTs separately (**Figure 4c**), we found that the more atrophic, A-type remnants were more frequently towards the tumor side of the rim; In contrast, early-atrophic A+BD PTs were enriched in the periphery of the rim (**Figures 4d and 4e**).

Importantly, we also observed PT remnants in the tumor center of desmoplastic metastases, and these occurred at a lower density than in the rim (**Supplementary Figures 2c and 2d**).

Together, these data suggested that the outer, “desmoplastic” rim represents early stages of fibrosis evolution, while the peritumoral stroma shows a higher degree of liver atrophy. The presence of PTs in the tumor center, including both A and A+BD types (**Supplementary Figures 2c and d**), provides compelling evidence that the metastatic stroma of desmoplastic CRLM is derived from adopted, remodeled liver parenchyma, which is easiest explained by previous replacement-type growth.

### Hepatic injury surrounds the desmoplastic rim

Inflammation invariably accompanies liver cell injury. For example, in non-alcoholic fatty liver disease (NAFLD), an increase in macrophages is a characteristic early sign of periportal damage, leading to fibrosis^26^. While the presence of lymphocytic cell infiltrates at the tumor-liver interface of the desmoplastic pattern has been demonstrated previously^3,27,28^, the spatial distribution of macrophages is not well investigated. In addition to immune cell infiltration, liver injury triggers the production of acute-phase proteins, such as C-reactive protein (CRP) and serum amyloid A1 (SAA1)/SAA2^29^, as well as increased expression of cytokeratin 18 (CK18)^30^. Recently, spatial transcriptomics of mouse liver has confirmed that injured hepatocytes expressing acute phase proteins form spatial clusters with macrophages, activated hepatic stellate cells expressing ASMA, and hepatobiliary progenitor cells^29^. In addition, fibrotic foci are characteristically accompanied by so-called ductular reactions, which represent nests of bile duct-like aggregates that are thought to be derived either directly from cholangiocytes, or via transdifferentiation from injured hepatocytes and induced progenitor cells^31^.

Our hypothesis that the desmoplastic rim represents a liver reparative reaction following hepatocyte-tumor cell injury predicts the presence of injury-related cellular constituents in and around the desmoplastic capsule. To test this, we generated spatial protein expression maps of the acute phase protein CRP and of CK18 across the entire desmoplastic rim and peritumoral liver, using digital quantitation of multiplex-immunohistochemistry (m-IHC). Tracing CRP and CK18 revealed a gradient of expression starting at the rim-liver interface **(Figures 5a–c**) and decreasing towards the perimetastatic liver parenchyma, demonstrating localized perimetastatic hepatocyte injury. Staining for cellular components of hepatic fibroinflammation with m-IHC, we found macrophage accumulation (cluster of differentiation [CD] 68 expression) and ductular reactions/bile duct remnants^26^ (CK7 expression, **Figures 5d–f**), consistent with an hepatic injury reaction that is most intense at the rim-liver interface.

**Figure 5:**
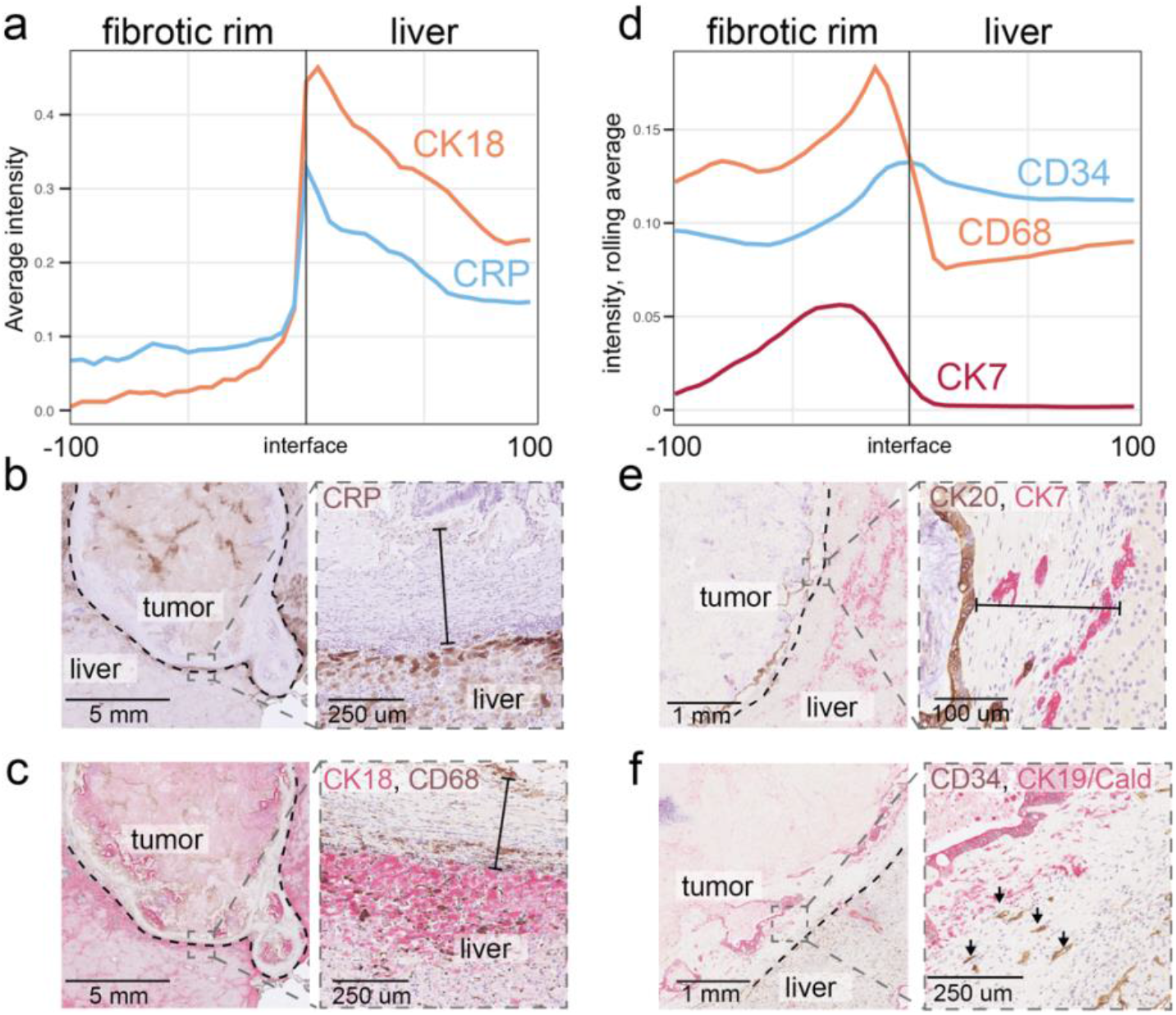
Protein gradients in the desmoplastic rim. **a)** Quantification of CRP and CK18 across the entire circumference of the fibrotic rim and in the perimetastatic liver parenchyma. Average values from n = 6 metastases, for which the entire desmoplastic rim was measured. **b)** and **c)** Representative examples of CRP, CK18/CD68 staining, as indicated. The dashed line marks the rim’s outer border; the solid line marks the rim’s thickness. **d)** Quantification of CD34 (endothelium), CD68 (macrophages), and CK7 (bile ducts and ductular reactions); rolling averages of results from n = 6 patients are shown. **e)** and **f)** Representative images of the indicated multiplex stains. A dashed line marks the outer border of the rim; the solid line marks the thickness of the rim, arrows point at CD34 vessel remnants in the rim.

### The perimetastatic capsule bears similarity to benign liver fibrosis

PTs are embedded in a specific stroma, positive for ASMA and low-affinity NGFR^2^. Therefore, we analyzed patterns of stromal protein expression from the liver parenchyma to the central tumor stroma via the fibrotic rim. We found zonation of the desmoplastic rim, such that expression of both NGFR and ASMA increased along the trajectory from the metastasis towards the liver (**Figures 6a–c**). Strikingly, NGFR expression was highest in the outer rim, where it resembled the profile of benign fibrotic conditions^32,33^. This prompted us to revisit stromal protein expression in livers with benign conditions. We found that the perilesional stroma in conditions such as focal nodular hyperplasia (FNH, **Figure 6d**) and cholangitis (**Figure 6e**) was NGFR^+^, similar to the outer part of the desmoplastic rim. Even in liver cirrhosis, we observed a similar NGFR^+^ stroma, which appeared to increase with a higher degree of liver plate atrophy (**Figure 6f–g**).

**Figure 6:**
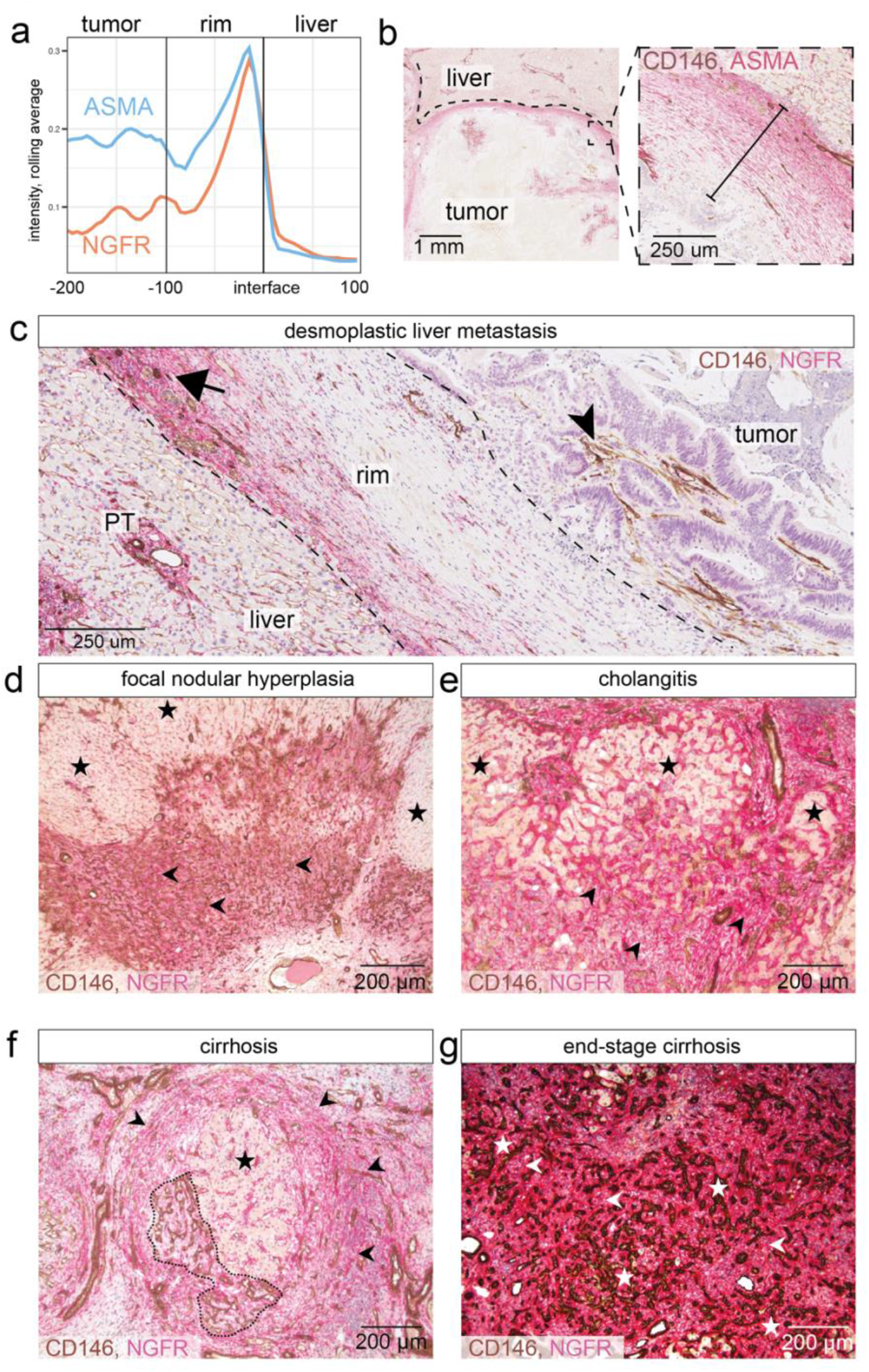
Similarity of the desmoplastic rim to benign fibrosis. **a)** Quantification of NGFR and ASMA protein expression in tumor, desmoplastic rim, and liver; rolling averages (n = 6, same as in Figure 5). Note increased expression of NGFR and ASMA towards outer, perihepatic edge of rim. **b)** Representative example of ASMA staining (red), with CD146 (brown) marking ductular reactions. **c**) Representative example of NGFR+ gradient; note strong NGFR expression (red) increasing towards liver. CD146 (brown); arrowhead: intratumoral stroma; arrow: portal tract remnant. **d)** to **g)** Staining for NGFR (stroma, red) and CD146 (ductular reaction, brown) for indicated diseases; **d) Focal nodular hyperplasia (FNH)**. Hepatocytes are delimited by fibrous tissue composed of NGFR+ stroma, containing prominent ductular reaction (CD146+). Stars: hepatocytes; arrowheads: ductular reaction. **e) Cholangitis**. Hepatocytes surround inflamed portal zone show atrophy with concomitant development of NGFR+ fibrosis. Stars: hepatocytes; arrowheads: fibrous stroma. **f) Liver cirrhosis**. hepatic tissue shows partial atrophy, a rim of NGFR+ stroma surrounds remnants of hepatocyte nodules, while ductular reaction (CD146+) has developed. Star: remnant of hepatic nodule; arrowheads: fibrous rim. Dotted region: ductular reaction. **g) Alcoholic steatohepatitis** associated with liver cirrhosis with intensive ductular reaction (CD146+), embedded in a strongly NGFR+ fibrous stroma. Stars: ductular reaction, arrowheads: fibrous stroma.

## Discussion

Despite their strong prognostic value, little is known about the biology of the histopathological growth patterns in liver metastases. The lack of clear associations between the patterns, known driver mutations and other genetic features such as MSI^15^ suggests unknown genetic traits as drivers of growth pattern biology. Intralesional growth pattern heterogeneity, on the other hand, could be explained by clonal differences, such that genetic or epigenetic changes facilitate either replacement or desmoplastic growth. This heterogeneity would largely remain undetected by clinical assessments of mutational status and MSI because these analyses are mainly based on bulk DNA sequencing (mutations) or do not regularly account for histopathological features at the required resolution (e.g., assessment of MSI heterogeneity by staining for mismatch repair proteins). A more likely explanation for the difficulties in connecting growth patterns to tumor biological traits is the multifactorial emergence of the patterns. In a multifactorial model, the growth patterns represent a unifying, convergent phenotype of diverse cellular traits; this would explain both their inter-patient and intra-lesional heterogeneity, as well as the fact that some replacement-type metastases convert to the desmoplastic type upon neoadjuvant chemotherapy, while others do not (**Figure 3** and ref. ^16^). As such, the growth patterns are potentially useful as predictive biomarkers because they biologically integrate different tumor and microenvironmental traits.

Our finding that the proportion of desmoplastic encapsulation determines prognosis in patients with CLRM (**Figure 2b–d**) provides support for the proposed multifactorial model, as it challenges previous data suggesting that the mere presence of replacement-type growth predicts a poor outcome, regardless of its extent^2^.

We, and others, have identified a group of patients whose metastases are completely encapsulated^2,15^. Both visual and extended digital scoring show that these patients have the most favorable prognoses (**Supplementary Figure 1**). However, the largest group of patients has a mixture of desmoplastic and replacement-type growth, and any clinicopathological sampling can only assess its composition to a limited extent. For these patients, extended scoring allows improved stratification. The prognosis of CRLM patients is strongly associated with relapse in the liver, and we suggest that the tumor’s ability to spread into the liver parenchyma - beyond the visible nodule that is surgically resected – is the main determinant for relapse. The growth pattern proportion-dependency of prognosis identified in this study is well in line with this interpretation.

The unifying feature of desmoplastic metastases is the lack of direct contact between the tumor cells and the surrounding hepatocytes. The origin of the rim that separates both compartments has so far been elusive^2,10,14^. Our findings show that the fibrotic rim is heterogeneous and represents a zonal evolution of distinct histological features starting from the liver-rim interface and extending to the rim-tumor border (**Figure 7**): Its outer (perihepatic) region closely resembles portal stroma (**Figures 6a - c**) and contains the most well-preserved liver parenchymal remnants, including A+BD PTs (**Figure 4d & e**), *ALB*^*+*^ -hepatocyte progeny (**Figure 5a**), and NGFR^+^ stroma (**Figure 6a**). In contrast, the inner rim, adjacent to the tumor, largely lacks A+BD PTs (**Figure 4d & e**), is devoid of hepatocyte remnants (**Figure 5a**) and progressively adopts a more intrametastatic stromal expression profile (**Figures 6a - c**). This suggests an evolutionary trajectory of the microenvironment from the outer zone of liver parenchymal damage (CD68^+^ macrophages, CK7^+^ ductular reactions, **Figure 5d**) towards the tumor center. Remarkably, these findings localize the zone of active formation of the rim to the liver side, which in some cases is several hundred micrometers away from detectable tumor cells. The presence of PTs in the center of desmoplastic metastases (**Supplementary Figure 2**) reveals previous spread into the hepatocyte compartments, and incorporation or co-option of the remaining stromal and PT architecture, lending further compelling support to a model of growth pattern plasticity, in this case from replacement growth (*“tumor wins”*) to desmoplastic encapsulation (*“liver wins”*). The opposite growth pattern conversion can also occur, and is the most likely explanation for the recently described escape phenomenon, where nodular outgrowths with a replacement-type pattern develop in otherwise largely desmoplastic-type metastases, usually after chemotherapy treatment^2^.

**Figure 7:**
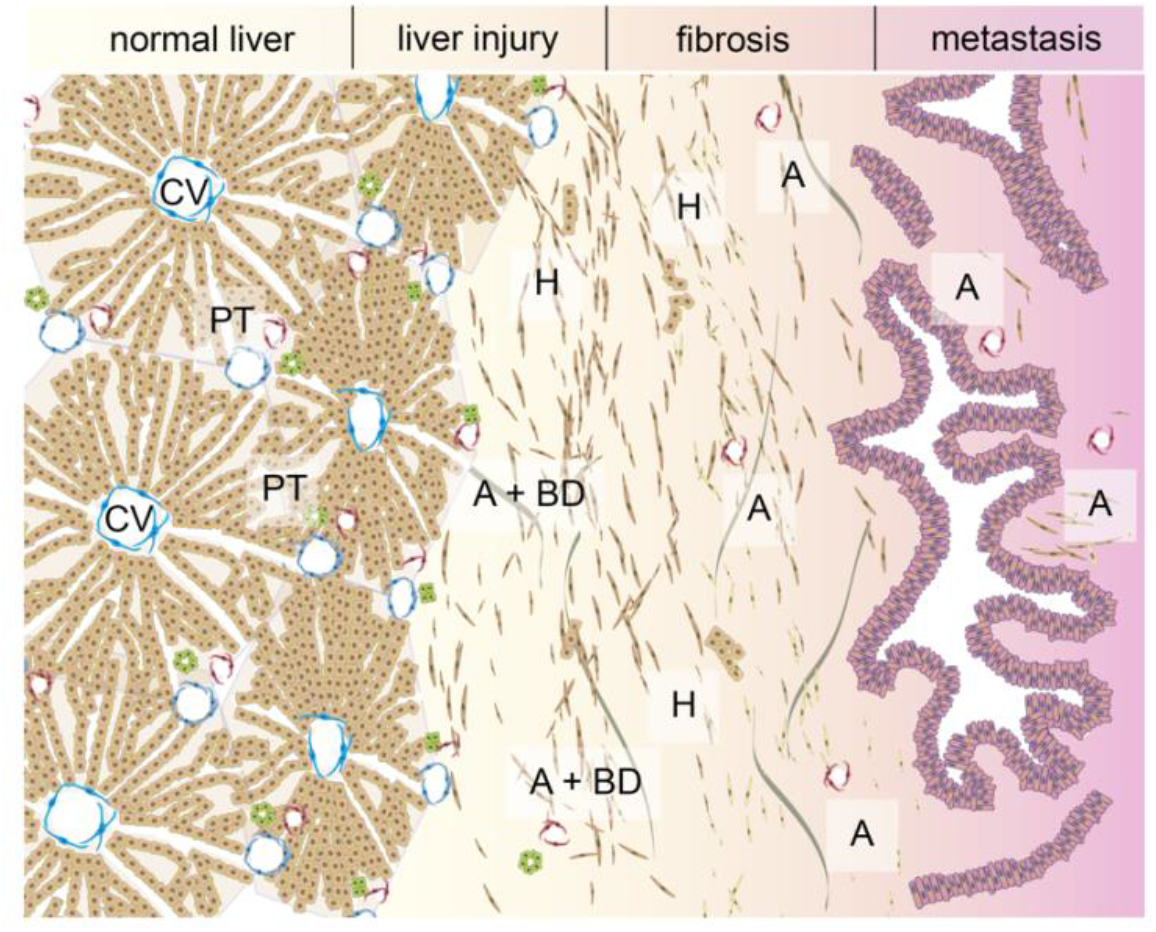
Illustration of zonation of the desmoplastic rim. Normal liver with central vein (CV) and portal tracts (PT) to the left. At the outer part of the rim, liver atrophy and fibrotic stroma similar to benign liver fibrosis can be seen. Portal tract remnants consisting of arteries and bile ducts (A+BD), only arteries, demarcated as previous PTs by portal stroma (A), and hepatocyte-derived cells can be found in the rim. The stroma in the tumor still contains those remnants although at lower frequency. Together, the histology of the rim and its clinical characteristics (potentially inducible by chemotherapy, and associated with decreased tumor cell viability, in conjunction with the proportion-dependency of outcome) suggest a new model in which the desmoplastic rim evolves upon failed invasion, and represents a completed hepatic injury reaction.

Together, our data support a model in which the desmoplastic rim primarily develops without active juxtracrine or paracrine signals from tumor cells, but rather as the combined result of liver injury and failed replacement-type growth. During temporal evolution, the tumor likely shapes its more central environment, which progressively diverges from the idiosyncratic stroma of the outer rim. From a tumor evolutionary perspective, we propose that replacement growth is the default pattern of most initial stages. If the replacement of hepatocytes becomes ineffective, or in case a sufficient liver-tumor cell damage reaction develops, the emerging fibrotic rim precludes further invasion, which explains the more favorable prognosis of patients with encapsulated CRLM (**Figures 2a & b**). In this model derived from our data, differences in the capability for replacement growth - for example, due to chemotherapy - can explain inter- and intralesional growth pattern heterogeneity. Alternatively, disturbances of blood supply and vascular remodeling connected to tumor growth could contribute to damage and atrophy of the peritumoral liver, and to the induction of encapsulating fibrosis. Finally, this model also accounts for the fact that encapsulated tumors are seen in some patients who have not received neoadjuvant chemotherapy: given a propitious balance of factors that favor a mature hepatic injury reaction *vs*. the tumor’s ability for replacement-growth, the latter stalls at a point where a sufficiently enveloping capsule can develop to prevent further replacement.

Our extended scoring requires multiple steps including extensive sampling of the invasion front, selecting slides to build an approximated central section, careful digital annotation, and expert review (**Figure 1b**). Given this complexity, and the fact that systematic sampling is part of the clinical routine at our Pathology Department, we cannot conclusively determine which specific steps in our approach are driving the difference between our findings and other recent data^2,14,15^. It is possible that the annotation resolution and/or the high number of individual sections per tumor nodule used in our dataset are needed to improve patient stratification. As extended scoring is prohibitingly time-consuming for clinical routine, developing image analysis support tools that leverage, e.g., machine learning may allow fine-grained prognostication based on the growth patterns in the future; standardized sampling, as suggested in the recent guidelines^2^, will then be particularly important. Meanwhile, we endorse the dichotomic use of 100% desmoplasia *vs*. all other fractions suggested in the scoring guidelines^2^ to identify those patients with an excellent prognosis after CRLM resection as a reproducible and clinically feasible approach.

Clinically, our findings nevertheless argue for a more granular growth pattern scoring than is currently recommended^2^, which can reasonably be achieved only by computer-aided image-analysis.

Biologically, our results highlight the role of tumor-hepatocyte-microenvironment interactions for driving metastasis invasion, and they suggest that targeting their crosstalk could elicit an hepatic injury reaction, promote perimetastatic fibrosis, and thereby provide an unexplored avenue for targeted treatment of liver metastases.

## Data Availability

All data produced in the present study are available upon reasonable request to the authors.

## Materials and Methods

### Patients

All consecutive patients with CRLM operated on between 2012 and 2015, were identified in electronic databases at Karolinska University Hospital, Huddinge, Sweden. All patients underwent surgical resection of one or more CRLM at Karolinska University Hospital. Previous or synchronous diagnosis of colorectal cancer was confirmed histologically. Clinical data were collected by retrospective review of the electronic patient records. Data on tumor mutations (such as *KRAS, BRAF, NRAS*, and MSI status) were extracted from routine clinical analyses. “Synchronous” were metastases diagnosed within three months of diagnosis of the primary tumor. “Neoadjuvant treatment” was defined as chemotherapy that was given within three months prior to metastasis surgery. “Right sided” was defined as cecum, ascending colon, hepatic flexure, and proximal two thirds of the transverse colon. “Left sided” was defined as the distal third of the transverse colon, the splenic flexure, sigmoid colon, descending colon, and the rectum. In case the precise location of a tumor in the transverse colon could not be determined, this tumor was considered right sided.

For IHC experiments, a series of n = 6 CRLM patients (n = 2 males and n = 4 females) was included. Of these, n = 3 metastases originated from left-sided primary colorectal adenocarcinomas, n = 3 from right-sided, and n = 3 patients received neoadjuvant chemotherapy. In n = 5 patients, margin-free tumor resection (pR0) was achieved. Tumors from n = 3 patients had confirmed *KRAS* mutations, two were *KRAS* wild type (one of which had an NRAS mutation), and for one, the *KRAS*/*BRAF*/*NRAS* mutation status was unknown. MSI status was unknown for all six tumors. For RNA ISH against *ALB*, four randomly selected metastases from these six were used.

Patients with benign fibrotic conditions and patients with cholangiocarcinoma were identified by the authors (CFM and BB) during routine clinical diagnosis, and are representative of histopathological findings in these cases. Antibody stains for these patients were part of clinical routine.

### Slide selection to approximate a panoramic central slice along the largest metastasis diameter

All H&E slides for each probe (liver resection specimen) were retrieved from the pathological archive and reviewed for each individual metastasis with an optical microscope. The slides approximating a panoramic central slice along the largest tumor diameter were selected for each individual metastasis. In cases where the growth pattern (GP) in non-central slices was significantly different from the GP in the selected slides (i.e., central and peripheral slices of a particular metastasis had different GPs), the latter were also included. To ensure consistency and quality in the selection process, all slides were reviewed by one experienced liver pathologist (CFM). Slides were digitized using a Hamamatsu NanoZoomer S360 digital slide scanner at 40x magnification.

### Digital annotation of the growth patterns

To annotate the GPs, the Hamamatsu software NDP.view 2, version 2.7.25, was used. Briefly, the entire invasion front of each slide was reviewed ensuring the quality of the WSIs before thoroughly evaluating and annotating the liver-tumor interface. Following the pre-determined GPs (desmoplastic, replacement type 1, replacement type 2 and pushing), a specific color and label was assigned to each one. Replacement types 1 and 2 were later combined as “replacement”, in line with the latest consensus guidelines and because of the difficulty in scoring them reproducibly.

Using the “Freehand Line” tool on NDP.view 2, the liver-tumor interface was manually annotated for each WSI following GP labelling. Adhering to GP scoring guidelines, capsule, portal zones and necrosis were not annotated, as they do not represent the liver-tumor interface^2^. After completing the annotation process, a Groovy script was developed to transfer the annotations to QuPath (version 0.2.3)^34^. In addition, the percentage of viable tumor cells within the tumor was estimated visually for each WSI.

### RNA *in situ* hybridization

RNA ISH was performed on 5 μm sections of formalin-fixed, paraffin-embedded patient samples using the RNAscope Multiplex Fluorescent Reagent Kit v2 (ACD, catalog no. 323100) according to the manufacturer’s instructions. The probe Hs-Alb (600941, C1) was used. Samples were baked at 60°C for 1h, deparaffinized and rehydrated followed by manual antigen target retrieval. Standard tissue pretreatment conditions were applied. Tyramide Signal Amplification (TSA®) Plus Fluorophores were diluted in RNAscope® Multiplex TSA buffer (ACD, catalog no. 322809) as follows: Fluorescein (Perkin Elmer, FP1168015UG diluted 1:1500) and Cy5 (Perkin Elmer, FP1171024UG, diluted 1:1000). Nuclear counterstain was performed with 4,6-diamindino-2-phenylindole (DAPI) immediately followed by mounting with ProLong Gold Antifade (P36930). Fluorescent images were taken with a Prime 95B sCMOS photometrics camera on a Nikon Eclipse Ti2 Inverted spinning disk microscope in 0.9 μm steps covering a total z range of 5 μm. A 2138.64 μm x 1527.6 μm field of view was captured. ImageJ, version 2.0.0, was used to generate maximum intensity projections of stacked images.

### Immunohistochemistry

Antibodies and specific staining conditions are specified in **Supplementary Table 1**. Briefly, formalin-fixed, paraffin-embedded (FFPE) tissue samples derived from routine pathological diagnostics of CRLM were cut at 4 μm thickness. Stains were performed on a Leica (Germany) BOND-MAX automated staining machine at Karolinska University Hospital, Huddinge, Sweden. Pretreatment was performed with Bond Epitope Retrieval Solution 2 EDTA (Leica) for 20 min. m-IHC stained slides were quantified as specified below for seven markers (NGFR, ASMA, CRP, CD68, CK18, CK7, CD34).

### Staining quantification on digitally scanned slides

Protein expression was quantified in the desmoplastic rim (DR), the perimetastatic liver parenchyma (PLP), and the tumor center (TC). The DR was annotated manually and defined as the area dominated by stromal cells facing the metastasis-liver interface outwards and tumor cells and/or necrosis inwards, thus yielding rims with different widths depending on the case. Next, the DR annotations were expanded at a fixed distance of 1500 μm into the perimetastatic liver parenchyma using a Groovy script. Then, all the portal tracts were manually annotated in the DR and PLP, subtracted from the DR and PLP annotations, and excluded from the quantification. Two cases were excluded from the quantification for the TC due to extensive tumor necrosis. As for the remaining cases (n = 4), a trained pixel classifier was used in one case to separate the stroma from the tumor tissue and to create tumor and stroma annotations due to complex and labor-intensive tumor/stroma separation, while the rest (n = 3) was annotated manually. Next, stain vectors for color deconvolution were determined in QuPath using the built-in tool for 3,3’ -Diaminobenzidine (DAB) and hematoxylin, and by selecting an area of pure staining for alkaline phosphatase (AP). Then, the stain vectors obtained were applied to all the images.

Next, each annotation, DR, PLP, and TC, was divided into 25 μm long square tiles. The mean intensity for each marker and distance to the invasion front were automatically calculated for each tile in QuPath. Finally, raw measurements were exported in tabular format for analysis.

### Statistical analysis of spatial annotations

Protein expression for each case and marker was analyzed in R. Because the thickness of the DR and TC varied for each case, the distances were normalized - including the PLP for consistency - such that the maximum width (= maximum distance) of each region corresponded to “100%”. Considering the liver-tumor interface as distance 0%, the full width of the PLP, DR, and TC regions were denoted as 100%, -100%, and -200%, respectively. Next, the distance of every tile from the liver-tumor interface was standardized per case, marker and region, using the following equation; standardized distance = 100/ (maximum distance * tile’s distance from the liver-tumor interface). For each case and marker, the mean intensities within the tiles were averaged over cut-off distances of 5% and over all regions, i.e. averaged over every 5% distance from -200% (TC) to 100% (PLP). Finally, the individual results were averaged for the whole series.

### Identification and annotation of the intratumoral portal tracts

All cases with an overall predominant desmoplastic GP (>95%, n = 55 patients) were selected for annotation of portal tracts. Since portal veins are most often obliterated, remnants of portal tracts were quantified either when hepatic arteries and bile ducts were seen together (artery and bile duct, “A+BD”) or when presented as isolated arteries surrounded by remnants of portal stroma (artery, “A”). Subsequently, all occurrences of A+BD and A throughout the tumor nodule were annotated on NDP.view 2 using the “pin” tool, while the DR area was annotated using the “freehand region” tool. For spatial quantification, two further annotations were added manually: an outer and inner border of the DR corresponding to the liver-rim interface and the rim-tumor interface, respectively. Next, all annotations were transferred from NDP.view 2 to QuPath using a Groovy script. After further refinement of the annotations, the minimal distance from each pin to both the outer (“facing the liver”) and inner (“towards the tumor”) borders of the rim was measured and data were exported from QuPath. The relative distance of each portaltract annotation to the liver-rim interface was finally calculated as follows: Relative distance to liver-rim interface = distance to liver-rim interface / (distance to liver-rim interface + distance to rim-tumor interface).

### Data analysis

Data were analyzed in R, version 2022.02.3. clinical table was built using gtsummary, version 1.6.1, and finalfit, version 1.0.4. Data visualization was done with ggplot2, version 3.3.6, as part of tidyverse, version 1.3.1. Survival analyses, both uni- and multivariate, were performed with survival and survminer, version 3.3-1 and 0.4.9, respectively.

## Acknowledgments

This study was supported by The Swedish Research Council (project nr. 2018-02023), The Swedish Society for Medical Research, the Åke Wiberg Foundation, the Jeansson Foundation, and the Karolinska Institute (all to MG). JE is supported by Region Stockholm and by the Bengt Ihre Foundation. NG is supported by the German Research Foundation. CFM is supported by The Swedish Society for Medical Research (PD21-0114). We are grateful for the support provided by the histological and immunohistochemical laboratories at the Department of Clinical Pathology and Cancer Diagnostics, Karolinska University Hospital, and Sólrún Kolbeinsdóttir for carefully checking the code used for the analysis of clinical data. We gratefully acknowledge the support of Peter Bankhead for staining quantification with QuPath. This study was in parts performed at the Live Cell Imaging core facility/Nikon Center of Excellence, at Karolinska Institutet, supported by the KI infrastructure council.

---

**Supplementary Figure 1:**
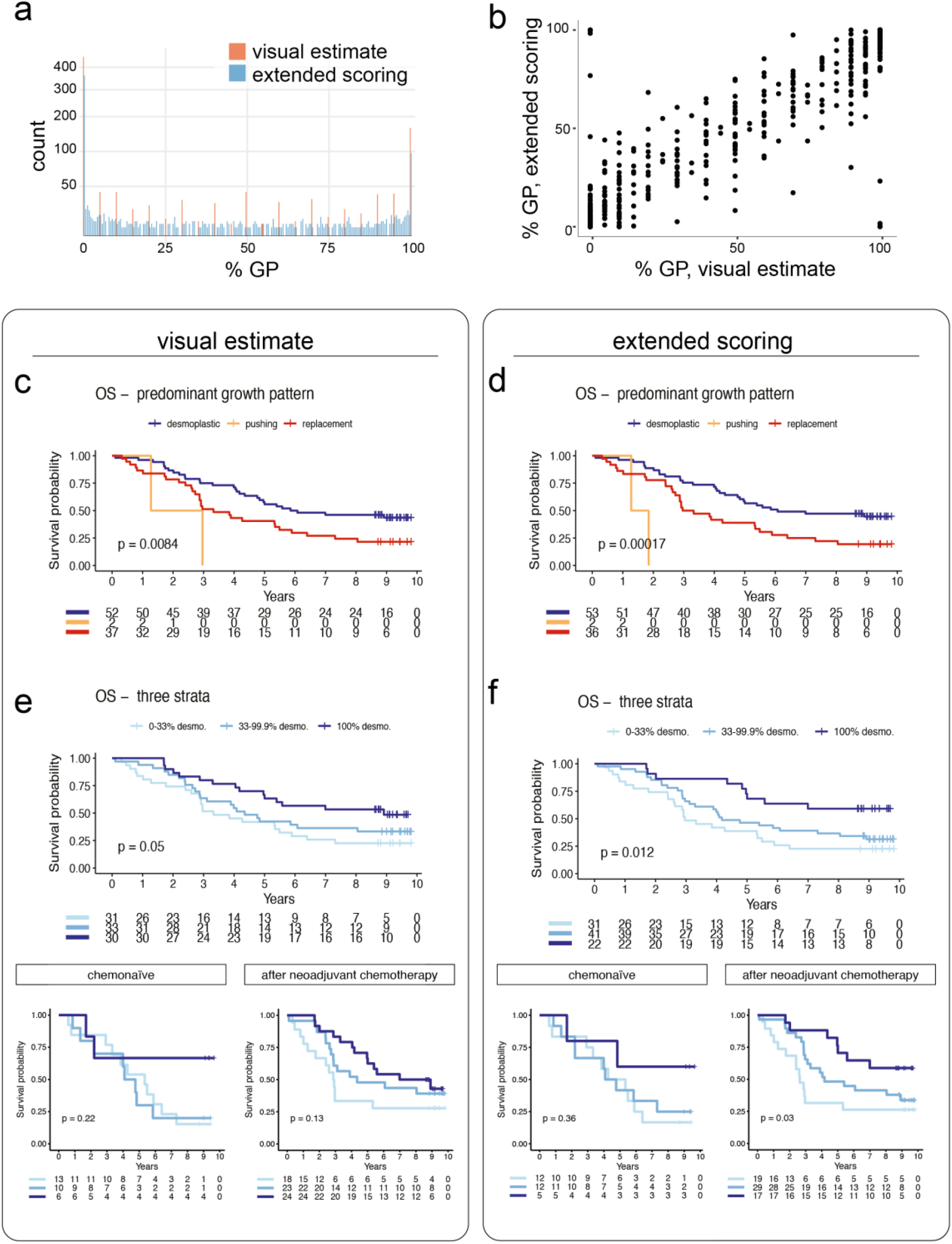
Visual estimates and extended scoring: distribution and prognostic value. **a)** Distribution of the results of visual estimates vs. digital annotations per slide (as percentage of any growth pattern per slide). **d)** Scatter plot of visual (horizontal) and digital (vertical) scorings of all growth patterns by the same raters (combined scores of two raters in [a] and [b]). **c – f)** Overall survival (OS) for the same patients when using visual scoring ([c] and [e]) or extended scoring ([d] and [f]) with the indicated stratifications. Log-rank p-values given in the plots.

**Supplementary Figure 2:**
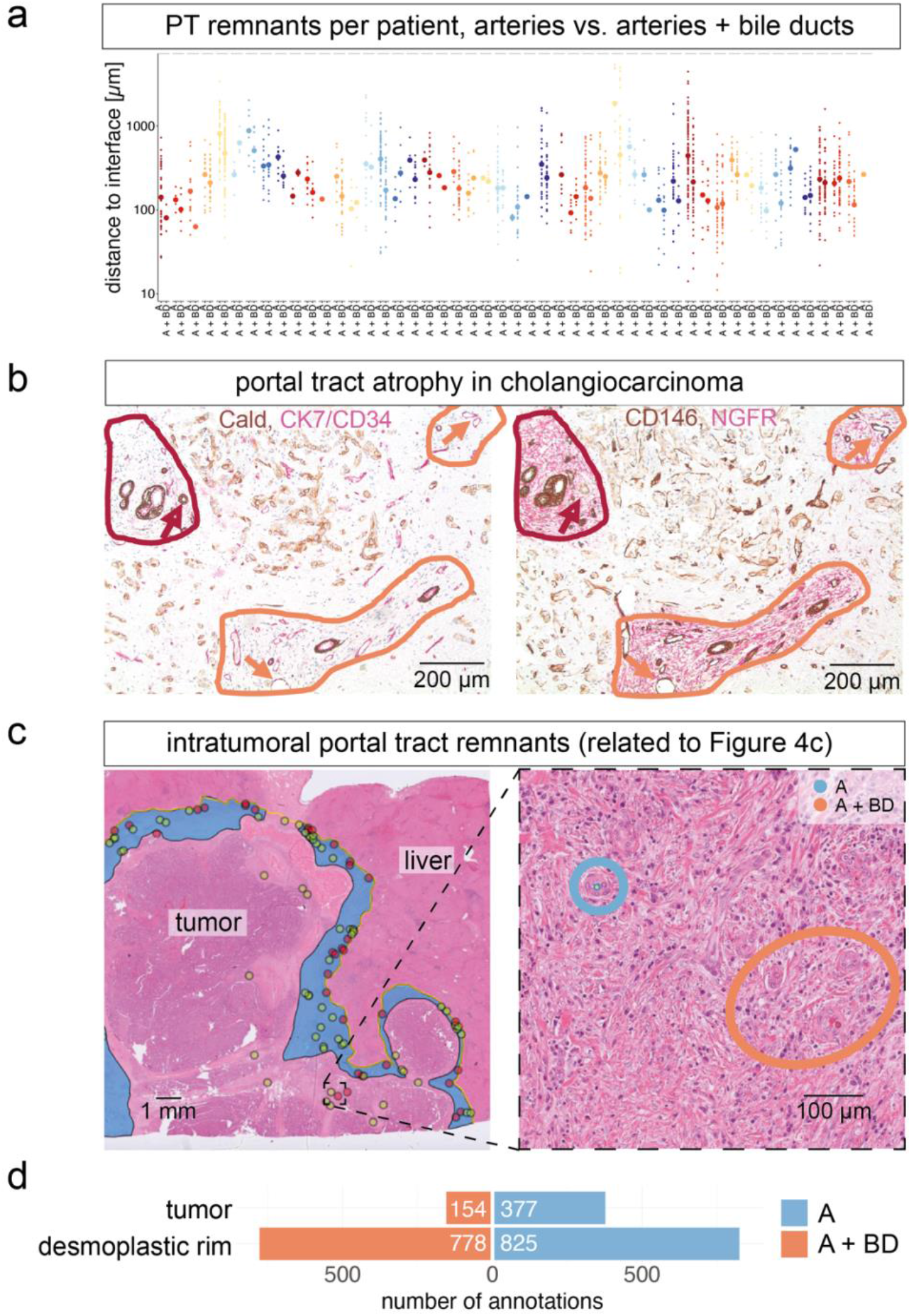
Quantification of liver parenchymal remnants. a) A and A+BD remnants per patient, different colors denot different patients (note, similar colors are used for different patients). **b)** Portal tract (PT) remnants in a cholangiocarcinoma, related to Figure 4b. Red: scant porta zones with preserved branches of the hepatic artery, bile duct (arrows), and variably portal stroma. Orange: remnants of porta zones with preserved branches of the hepatic artery and variably portal stroma. Preserved branches of the portal vein can still be detected (orange arrows). **c)** Whole-slide image (WSI) annotated for the desmoplastic rim and portal tracts as in Figure 4c: yellow line marks border of the desmoplastic rim and the liver; blue area marks the rim; red dots mark PTs consisting of artery and bile duct (A+BD), green dots mark PTs consisting of artery remnants (A). **d)** Quantification of the distribution of intratumoral PT remnants compared to the PT remnants in the rim; absolute numbers are given, without correction for the area. Cald: Caldesmon. CK7: cytokeratin 7, CD34: cluster of differentiation 34.

