## Supplementary Table 1 for "An idiosyncratic, zonated stroma encapsulates desmoplastic liver metastases and originates from injured liver"

**Supplementary Table 1. List of antibodies for multiplex-immunohistochemistry.** *CRP stains were done using a Ventana BenchMark Ultra autostainer, all other stains as described in *Methods*

| *Antibody* | *Clone* | *Dilution* | *Product code* |
|---|---|---|---|
| *Actin (smooth muscle)* | 1A4 | 1:500 | DAKO M0851 |
| *Caldesmon* | h-CD | 1:300 | DAKO M3557 |
| *CD34* | QBEnd/10 | 1:50 | DAKO M7165 |
| *CD68* | PG-M1 | 1:100 | DAKO M0876 |
| *CK18* | DC-10 | 1:50 | DAKO M7010 |
| *CK7* | RN7 | 1:200 | NCL-L-CK7-560 |
| *CD146* | UMAB154 | 1:200 | Origene-UM800051 |
| *NGFR* | polyclonal | 1:500 | Atlas-HPA004765 |
| *CRP* | Y284 | 1:200 | Abcam-ab271830* |
